# Comparative Effectiveness of Ticagrelor vs. Prasugrel in Patients with Acute Coronary Syndrome Undergoing Percutaneous Coronary Intervention

**DOI:** 10.64898/2026.08.13.26360416

**Authors:** Chang Hoon Han, Anna Ostropolets, Clair Blacketer, Christophe G. Lambert, Ben S. Gerber, Jose D. Posada, Farnoosh H. Sheikhi, Justin M. Petucci, Thamir M Alshammari, Marc A. Suchard, Michael E. Matheny, Christianus H. Setiawan, Mereeja Varghese, Aamirah Vadsariya, Syed Muhammad Musa Ali Rizvi, Behnood Bikdeli, Seng Chan You

## Abstract

**Background:** Ticagrelor and prasugrel are recommended P2Y₁₂ inhibitors for patients with acute coronary syndrome (ACS) undergoing percutaneous coronary intervention (PCI), yet uncertainty persists regarding their direct comparative evidence and guideline recommendations differ.

**Methods:** We conducted a multinational retrospective new-user cohort study across 7 claims and electronic health record databases. Adults with ACS undergoing first PCI who initiated ticagrelor or prasugrel were included; patients with prior major ischemic or hemorrhagic events or oral anticoagulant use were excluded. The primary outcome was 1-year major adverse cardiovascular events (MACE: all-cause mortality, acute myocardial infarction, or stroke). Secondary outcomes included net adverse clinical events (NACE) and individual components. Propensity scores were estimated using large-scale L1-regularized logistic regression and applied through stratification. Prespecified diagnostics (covariate balance, empirical equipoise, and systematic error) determined eligibility of each database for inclusion in meta-analysis. Database-specific hazard ratios (HRs) were combined using Bayesian random-effects meta-analysis.

**Results:** Among 7 participating databases, 3 met prespecified diagnostic criteria and were included in the primary meta-analysis, comprising 133,718 patients from one nationwide Korean claims database and two U.S. commercial claims databases (ticagrelor, 109,639; prasugrel, 24,079). For 1-year MACE, the pooled HR for ticagrelor versus prasugrel was 1.28 (95% credible interval [CrI], 0.89–1.88), with substantial between-database heterogeneity. Sensitivity analyses across alternative time-at-risk definitions and propensity score matching were consistent. No statistically credible differences were observed for NACE (HR 1.23, CrI 0.88–1.75), all-cause mortality (HR 1.17, CrI 0.78–1.77), cardiovascular mortality (HR 1.23, CrI 0.81–1.87), ischemic events (HR 1.28, CrI 0.88–1.90), hemorrhagic events (HR 1.01, CrI 0.72–1.39), acute myocardial infarction (HR 1.30, CrI 0.88–1.94), stroke (HR 1.09, CrI 0.73– 1.58), or gastrointestinal bleeding (HR 1.04, CrI 0.77–1.41). In a post hoc meta-analysis restricted to the two U.S. databases, the pooled HR for 1-year MACE was 1.49 (95% CrI 1.05–2.10).

**Conclusions:** In this pre-specified multinational observational study, no statistically credible difference in 1-year MACE was observed between ticagrelor and prasugrel in patients with ACS undergoing PCI. However, substantial cross-database heterogeneity warrants further investigation into context-specific comparative effectiveness and safety.

**CLINICAL PERSPECTIVE:** *What is new?:*

- In a multinational new-user active comparator study across one nationwide Korean claims database and two U.S. commercial claims databases, ticagrelor and prasugrel did not show a statistically credible overall difference in 1-year MACE after PCI for ACS.
- Despite a prespecified protocol, standardized execution, and empirical calibration, substantial cross-database heterogeneity was observed, with neutral estimates in Korea and prasugrel-favoring ischemic signals in U.S. databases.

*What are the clinical implications?:*

- These findings do not support superiority of ticagrelor or prasugrel for 1-year MACE across all health-care settings.
- Substantial cross-database heterogeneity argues against assuming that the comparative effectiveness and safety of ticagrelor and prasugrel are uniform across populations and care settings.
- Selection between ticagrelor and prasugrel should remain individualized, integrating individual patient risk profiles and local care considerations rather than presume a universally preferred agent.

## INTRODUCTION

Dual antiplatelet therapy (DAPT) with aspirin plus a P2Y12 receptor inhibitor is a cornerstone of treatment for patients with acute coronary syndrome (ACS) undergoing percutaneous coronary intervention (PCI).^1,2^ In contemporary practice, ticagrelor and prasugrel are generally preferred over clopidogrel, supported by pivotal trials and reinforced by guideline recommendations.^1–4^

However, direct randomized comparisons of ticagrelor and prasugrel remain limited in number and heterogeneous in study population and design. PRAGUE-18, which compared prasugrel and ticagrelor in patients with acute myocardial infarction (AMI) undergoing PCI, did not show a significant difference between agents for its net clinical endpoint, but the interpretation was limited by premature termination and inadequate statistical power.^5,6^ In contrast, ISAR-REACT 5 reported a lower incidence of the composite endpoint (death, myocardial infarction, or stroke) with prasugrel than with ticagrelor, without a statistically significant increase in major bleeding.^7^ This result challenged earlier interpretations that had favored ticagrelor in ACS and sparked debate about trial design, interpretation, and generalizability.^8–10^ More recently, TUXEDO-2 failed to demonstrate noninferiority of ticagrelor to prasugrel among patients with diabetes and multivessel coronary artery disease undergoing PCI, extending the randomized evidence to a selected high-risk population.^11^

Guideline and consensus recommendations have not fully converged in their interpretation of the available comparative evidence. The European Society of Cardiology (ESC) introduced a qualified preference for prasugrel over ticagrelor in patients with ACS proceeding to PCI, largely reflecting the results of ISAR-REACT 5.1,^12^ In contrast, contemporary U.S. guidelines from the American College of Cardiology (ACC) and American

Heart Association (AHA) continue to recommend either agent without preference, reflecting ongoing uncertainty.^2,13^ More recently, however, a 2026 ACC Scientific Statement acknowledged that some comparative data favor prasugrel and stated that choosing prasugrel over ticagrelor is reasonable in patients without contraindications.^14^

While a shift toward prasugrel has occurred in some settings following ISAR-REACT 5, ticagrelor has remained substantially more commonly used than prasugrel in the United States and Korea.^15–18^ Such variation in adoption across regions and populations is consistent with persistent uncertainty regarding the optimal P2Y12 inhibitor. Several observational comparative-effectiveness studies have evaluated ticagrelor and prasugrel in routine clinical practice, but their mixed findings remain subject to methodological concerns inherent to observational comparisons, including potential residual confounding, group comparability, and limited generalizability.^18–22^ To address these limitations, we conducted a large-scale, multinational observational cohort study within the Observational Health Data Sciences and Informatics (OHDSI) Evidence Network using standardized data, a prespecified protocol, and validated analytic methods across multiple databases.^23,24^

## METHODS

We performed a comparative new-user cohort study of ticagrelor versus prasugrel among patients with acute coronary syndrome (ACS) undergoing percutaneous coronary intervention (PCI), using routinely collected electronic health record and administrative claims data standardized to the Observational Medical Outcomes Partnership Common Data Model (OMOP-CDM).^25^

The study included 7 claims-based and electronic health record (EHR) databases from the United States and South Korea. Claims data sources included the Merative™ MarketScan® Commercial Claims and Encounters (CCAE), Merative™ MarketScan® Medicare Supplemental and Coordination of Benefits (MDCR), and HealthVerity databases in the United States and the nationwide Health Insurance Review and Assessment (HIRA) database in South Korea. EHR data sources included UMass Memorial Health, Penn State Health, and Stanford Health Care. Additional details are provided in eMethod 1.

Analyses were executed in a distributed manner, with patient-level data retained within each center’s local environment and only aggregated results shared centrally for interpretation and meta-analysis. This federated study framework has been previously described and validated for large-scale comparative effectiveness and safety research.^26,27^

All analyses followed a prespecified analytic protocol registered in the Heads of Medicines Agencies–European Medicines Agency Real-World Data catalogue.^28^ This study was approved by the institutional review boards of Yonsei University Health System (4-2024-0718), UMass Chan Medical School (MOD00007080), and the University of New Mexico Health Sciences Center (26-282). This study was reported according to the Strengthening the Reporting of Observational Studies in Epidemiology (STROBE) reporting guideline.

### Study Cohorts and Exposure

The study population consisted of adult patients (≥18 years) with ACS who underwent PCI and initiated either ticagrelor or prasugrel. The index date was the date of the first PCI. To ensure adequate characterization of baseline covariates and to minimize left censoring, patients were required to have at least 365 days of continuous observation prior to the index date.

Treatment exposure was defined as initiation of ticagrelor or prasugrel from 1 day before through the index date. We excluded patients exposed to the alternative P2Y₁₂ inhibitor, warfarin, or direct oral anticoagulants during the 6-month pre-index period, as well as those with any prior history of stroke or gastrointestinal bleeding. Detailed cohort definitions and study design are described in eMethod 2.29,30

### Study Outcomes and Time-at-Risks

The primary outcome was 1-year major adverse cardiovascular events (MACE), defined as a composite of all-cause mortality, acute myocardial infarction (AMI), and stroke. Secondary outcomes included net adverse clinical events (NACE), defined as a composite of all-cause mortality, AMI, stroke, and GI bleeding; all-cause mortality; cardiovascular mortality, operationally defined as a death event accompanied by a record of sudden cardiac death, AMI, ischemic or hemorrhagic stroke, or hospitalization for heart failure within the 30 days preceding or on the date of death; ischemic events, defined as a composite of AMI and ischemic stroke; hemorrhagic events, defined as a composite of hemorrhagic stroke and GI bleeding; all stroke; and the individual components of the composite outcomes, including AMI, ischemic stroke, hemorrhagic stroke, and GI bleeding.

All outcomes were defined using prespecified, code-based algorithms implemented in the OMOP-CDM, with diagnosis concepts derived primarily from International Classification of Diseases, Tenth Revision codes mapped to OMOP Standardized Vocabularies.^31^ Individual outcomes were validated through targeted chart review in the Yonsei University Health System (YUHS) database. Detailed definitions of individual outcomes, as well as the procedures and results of chart-based validation, are provided in eMethod 3.^32,33^

The primary time-at-risk was 1 year following cohort entry. Prespecified sensitivity analyses evaluated fixed 1-and 3-month windows and an as-treated definition. Detailed follow-up timelines and time-at-risk definitions are provided in eMethod 2.

### Statistical Analysis and Diagnostics

To address potential confounding between treatment groups, propensity score (PS) adjustment of baseline covariates was applied in all analyses. PSs were estimated separately within each data source using large-scale L1-regularized logistic regression.^34,35^ Covariates included demographic characteristics, medical conditions, medication use, procedures, devices, and measurements assessed over 7- and 365-day pre-index windows. Race and ethnicity were included when available. Covariate balance before and after PS adjustment was assessed using absolute standardized mean differences (aSMDs). The primary analysis used PS stratification into five strata, selected to retain the full study population and statistical efficiency given unequal cohort sizes. As a sensitivity analysis, 1-to-1 PS matching was performed to evaluate the robustness under an alternative adjustment strategy. Within each database, incidence rates were calculated, and Cox proportional hazards models estimated hazard ratios (HRs) with 95% confidence intervals (CIs) comparing ticagrelor with prasugrel. PS-stratified Kaplan-Meier curves were generated to visualize adjusted event-free survival probabilities.^36^

To quantify systematic error and support empirical calibration, 100 negative control outcomes not plausibly related to either treatment or study outcomes were analyzed using the same analytic pipeline.37,^38^ (eMethod 4) Prespecified study diagnostics required assessment of covariate balance after PS adjustment, evaluation of empirical equipoise based on the overlap of preference score distributions between treatment groups, and quantification of systematic error using negative control outcomes to calculate the expected absolute systematic error (EASE). Only results that met all diagnostic thresholds, adequate balance (maximum aSMD <0.1) of pre-selected covariates (eTable 1) after PS adjustment, sufficient empirical equipoise (≥20% of patients with preference scores between 0.3 and 0.7), and acceptable systematic error (EASE <0.25), were included in the final evidence synthesis.33,^39^ Database-specific HRs and 95% CIs were empirically calibrated; all figures and tables present calibrated estimates unless otherwise specified.

For outcomes meeting all diagnostic criteria, database-specific HRs were combined using Bayesian random-effects meta-analysis on the log-HR scale. A weakly informative normal prior was specified for the overall mean log HR (μ ∼ normal(0, 10²)), and a weakly informative half-normal prior was specified for the between-database heterogeneity parameter (τ ∼ half-normal(0.30)).40

Study packages were developed using R versions compatible with local data partner environments, including R versions in the 4.2–4.4 series, with analytic code publicly available in the Ticagrelor vs. Prasugrel repositories.41,^42^ Final aggregation of database-specific results, meta-analysis, and result organization were performed using R version 4.4.3.

### Post Hoc Exploratory Analyses

We conducted a post hoc exploratory meta-analysis restricted to U.S. data sources to assess the consistency of findings within the U.S. healthcare context. The same cohort definitions, statistical models, and diagnostic criteria were applied.

Furthermore, in a separate analysis, we addressed incomplete capture of inpatient medication prescriptions in CCAE and MDCR by applying an expanded cohort definition to these two databases. Specifically, the treatment exposure ascertainment window was extended from day −1 through day 0 to day −1 through day +1 relative to PCI. Cohort definitions in the other databases, as well as all other analytic procedures and prespecified diagnostic criteria, were retained for the meta-analysis. The rationale and detailed methods are provided in eMethod 5.

## RESULTS

### Study population and characteristics

Across 7 participating databases, application of prespecified study diagnostics resulted in inclusion of 3 databases—HIRA, HealthVerity, and CCAE—in the primary meta-analysis, whereas MDCR, UMMH, SHC, and PSH were excluded (Table 1). Empirical equipoise for each participating database is illustrated by the preference score distributions in eFigure 2. Covariate balance for prespecified diagnostic variables in the included databases is summarized in Table 2 and eTables 2–3, and covariate balance before and after PS stratification for all participating databases is shown in eFigures 3–4. The distribution of negative control outcome effect size estimates used to calculate the EASE is presented in eFigures 5–6. A detailed study flowchart is provided in eFigure 1.

**Table 1.** Database-level study diagnostics and meta-analysis inclusion.

| Database* | Ticagrelor<br>(N) | Prasugrel<br>(N) | Diagnostics† |  |  | Inclusion in<br>Meta-analysis* |
| --- | --- | --- | --- | --- | --- | --- |
| | | | Max<br>aSMD<br>($<0.1$) | Empirical<br>equipoise<br>($\geq 20\%$) | EASE<br>($<0.25$) | |
| **HIRA** | 90101 | 15919 | **0.07** | **39.1%** | **0.02** | **Included** |
| **HealthVerity** | 16177 | 5586 | **0.05** | **78.3%** | **0.06** | **Included** |
| **CCAE** | 3361 | 2574 | **0.06** | **87.0%** | **0.13** | **Included** |
| MDCR | 982 | 508 | 0.18 | **81.0%** | **0.10** | Not included |
| UMMH | 1164 | 189 | 0.64 | **53.2%** | 0.58 | Not included |
| SHC | 684 | 47 | 0.69 | 18.5% | 0.83 | Not included |
| PSH | 135 | 25 | 0.47 | 17.1% | 0.75 | Not included |

\*Databases meeting all three prespecified diagnostic criteria were included in the meta-analysis and are shown in bold.
† Maximum aSMD assessed residual covariate imbalance after propensity score adjustment; empirical equipoise assessed overlap in treatment preference between groups; and EASE quantified expected systematic error using negative control outcomes. Values meeting the prespecified diagnostic thresholds are shown in bold.
Abbreviations: aSMD, absolute standardized mean difference; EASE, expected absolute systematic error; HIRA, Health Insurance Review and Assessment; CCAE, Commercial Claims and Encounters; MDCR, Medicare Supplemental and Coordination of Benefits; UMMH, UMass Memorial Health; SHC, Stanford Health Care; PSH, Penn State Health.

**Table 2.** Baseline characteristics of included patients from Health Insurance Review and Assessment database.

|  | Before PS stratification |  |  | After PS stratification |  |  |
| --- | --- | --- | --- | --- | --- | --- |
|  | Ticagrelor<br>(N= 90101) | Prasugrel<br>(N= 15919) | aSMD | Ticagrelor<br>(N= 90101) | Prasugrel<br>(N= 15919) | aSMD |
| Age group |  |  |  |  |  |  |
| 25-29 | 0.1 | 0.2 | 0.009 | 0.1 | 0.3 | 0.023 |
| 30-34 | 0.5 | 0.6 | 0.014 | 0.6 | 0.5 | 0.003 |
| 35-39 | 1.9 | 2.0 | 0.008 | 1.9 | 2.0 | 0.008 |
| 40-44 | 4.3 | 5.2 | 0.040 | 4.5 | 4.5 | 0.001 |
| 45-49 | 8.6 | 9.8 | 0.043 | 8.9 | 9.4 | 0.018 |
| 50-54 | 12.6 | 14.7 | 0.060 | 13.1 | 13.3 | 0.008 |
| 55-59 | 15.9 | 19.1 | 0.085 | 16.5 | 16.9 | 0.010 |
| 60-64 | 16.1 | 20.1 | 0.103 | 16.8 | 17.2 | 0.011 |
| 65-69 | 13.1 | 15.5 | 0.068 | 13.4 | 13.1 | 0.010 |
| 70-74 | 10.3 | 8.9 | 0.049 | 9.8 | 9.9 | 0.003 |
| 75-79 | 8.4 | 2.7 | 0.252 | 7.5 | 6.9 | 0.024 |
| 80-84 | 0.1 | 0.0 | 0.250 | 0.0 | 0.0 | 0.004 |
| 85-89 | 2.2 | 0.2 | 0.186 | 1.8 | 1.0 | 0.065 |
| Sex |  |  |  |  |  |  |
| Female | 19.3 | 12.3 | 0.192 | 18.1 | 18.2 | 0.003 |
| Male | 80.7 | 87.7 | 0.192 | 81.9 | 81.8 | 0.003 |
| Comorbidities* |  |  |  |  |  |  |
| Essential hypertension | 70.6 | 70.5 | 0.003 | 70.7 | 71.4 | 0.016 |
| Diabetes mellitus | 41.7 | 39.5 | 0.043 | 41.3 | 41.7 | 0.007 |
| Hyperlipidemia | 84.0 | 85.6 | 0.043 | 84.4 | 85.7 | 0.036 |
| Obesity | 1.9 | 1.7 | 0.013 | 1.8 | 1.4 | 0.037 |
| Renal impairment | 6.5 | 4.1 | 0.107 | 6.0 | 5.8 | 0.007 |
| Atrial fibrillation | 2.1 | 1.8 | 0.023 | 2.1 | 2.2 | 0.007 |
| Heart failure | 26.3 | 23.5 | 0.065 | 25.9 | 25.5 | 0.008 |
| Peripheral vascular disease | 18.0 | 18.5 | 0.015 | 17.9 | 17.0 | 0.024 |
| Medication use† |  |  |  |  |  |  |
| Aspirin | 98.9 | 98.6 | 0.029 | 98.9 | 98.9 | 0.002 |
| Abciximab | 10.5 | 7.7 | 0.097 | 10.0 | 10.3 | 0.010 |
| Statins | 91.9 | 87.0 | 0.159 | 91.9 | 91.4 | 0.018 |
| Beta blockers | 74.9 | 72.9 | 0.045 | 75.7 | 73.7 | 0.045 |
| Calcium channel blockers | 45.8 | 47.6 | 0.037 | 46.1 | 48.4 | 0.046 |
| ACE inhibitors | 30.6 | 23.5 | 0.161 | 30.3 | 31.2 | 0.020 |
| Angiotensin II receptor blockers | 47.2 | 51.4 | 0.083 | 48.1 | 47.8 | 0.005 |
| Proton pump inhibitors | 58.7 | 53.9 | 0.099 | 58.3 | 58.7 | 0.007 |
| Diuretics | 36.5 | 27.4 | 0.197 | 35.0 | 34.8 | 0.003 |
| Insulins and analogues | 14.9 | 11.2 | 0.110 | 14.2 | 14.2 | 0.001 |
| Antidiabetic drugs, except insulins | 26.8 | 28.0 | 0.026 | 27.0 | 28.1 | 0.025 |

Race data was not covered in the database. Values are percentages unless otherwise indicated.
\*Defined as recorded diagnosis within 365 days before index date.
†Defined as recorded prescription within 7 days before index date.
Abbreviations: PS, propensity score; aSMD, absolute standardized mean difference; ACE, angiotensin-converting enzyme.

Among included databases, the analytic cohort comprised 109,639 ticagrelor users and 24,079 prasugrel users: HIRA (90,101 vs 15,919), HealthVerity (16,177 vs 5,586), and CCAE (3,361 vs 2,574). Baseline characteristics of patients in the included databases are summarized in Table 2 (HIRA) and eTables 2–3 (HealthVerity and CCAE). Race data were unavailable in HIRA and CCAE and incompletely captured in HealthVerity, where most patients with recorded race were White.

For the primary 1-year analysis, median (IQR) follow-up was 365 (365–365) days in both exposure groups in HIRA; 365 (281–365) days for ticagrelor and 365 (276–365) days for prasugrel in HealthVerity; and 365 (147–365) days and 365 (209–365) days, respectively, in CCAE (eTable 4). Incidence rates per 1,000 person-years for all outcomes of interest, along with minimum detectable relative risks, are reported in eTable 5.

### Primary outcome

For the primary endpoint of 1-year MACE, database-specific HRs are shown in Figure 1. In HIRA, there was no significant difference between ticagrelor and prasugrel (HR, 1.01; 95% CI, 0.93–1.10). In contrast, both US databases demonstrated higher hazards of MACE with ticagrelor compared with prasugrel: HR 1.50 (95% CI, 1.24–1.81) in HealthVerity and HR 1.47 (95% CI, 1.17–1.85) in CCAE (Figure 1). Propensity score-adjusted Kaplan-Meier curves of individual databases are shown in Figure 2.

**Figure 1.**
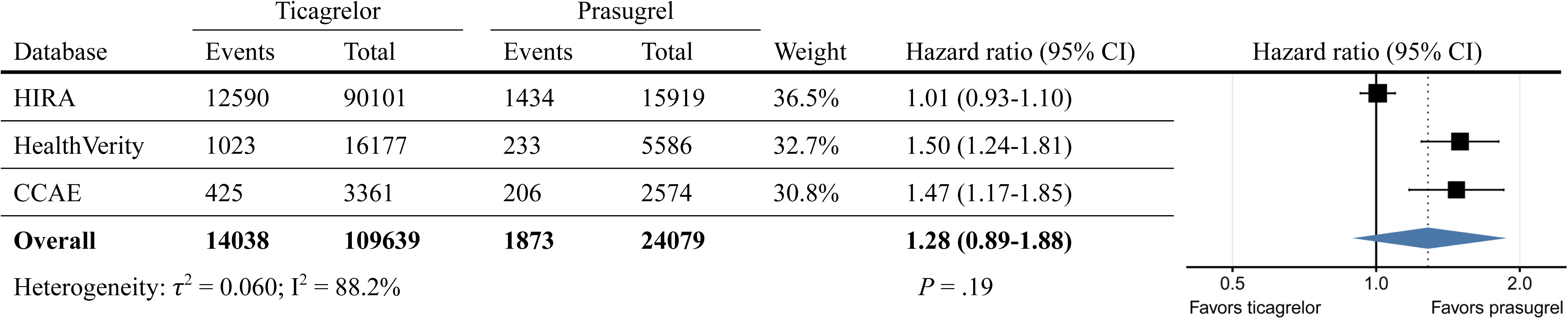
Comparative effectiveness of ticagrelor versus prasugrel for 1-year major adverse cardiovascular events. Forest plot of calibrated hazard ratios (HRs) for 1-year major adverse cardiovascular events (MACE), defined as a composite of all-cause mortality, acute myocardial infarction, and stroke (ischemic or hemorrhagic), comparing ticagrelor with prasugrel in patients with acute coronary syndrome undergoing percutaneous coronary intervention. Database-specific HRs and 95% confidence intervals (CIs) were estimated using propensity score-stratified Cox proportional hazards models, and were empirically calibrated. The pooled HR was obtained using Bayesian random-effects meta-analysis and is presented with its 95% credible interval (CrI). Weights are normalized random-effects inverse-variance weights based on the posterior median estimate of between-database heterogeneity. Square sizes are proportional to the weights, and horizontal lines indicate 95% CIs for database-specific estimates. The diamond represents the pooled posterior mean HR with its 95% CrI. The vertical solid line indicates no difference (HR = 1.0). HRs greater than 1.0 favor prasugrel over ticagrelor. Between-database heterogeneity is summarized by τ², the estimated between-database variance in the true log HRs, and I², the proportion of total variability attributable to between-database heterogeneity. *P* represents the P value for the overall pooled effect. *The pooled estimate is derived from Bayesian random-effects meta-analysis; the interval shown represents a 95% credible interval (CrI). Abbreviations: CI, confidence interval; HIRA, Health Insurance Review & Assessment Service; CCAE, Merative™ MarketScan® Commercial Claims and Encounters database.

**Figure 2.**
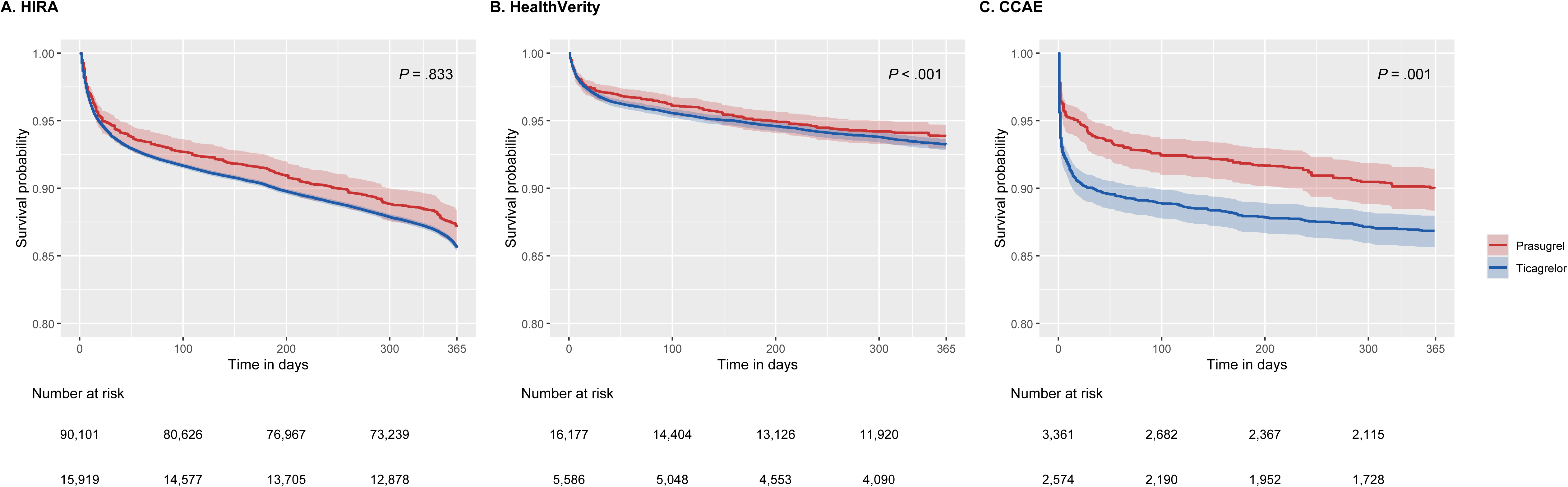
Propensity score-adjusted Kaplan-Meier curves for 1-year major adverse cardiovascular events. Propensity score-adjusted Kaplan-Meier curves comparing ticagrelor and prasugrel in patients with acute coronary syndrome undergoing percutaneous coronary intervention in (A) HIRA, (B) HealthVerity, and (C) CCAE databases. Adjustment was achieved using propensity score stratification.^36^ Survival probabilities represent event-free survival for the primary outcome of 1-year major adverse cardiovascular events (MACE). Shaded areas indicate 95% confidence intervals (CIs). P values correspond to propensity score–stratified Cox proportional hazards models. Numbers at risk at prespecified time points are shown below each panel. Abbreviations: CI, confidence interval; HIRA, Health Insurance Review & Assessment Service; CCAE, Merative™ MarketScan® Commercial Claims and Encounters database.

Negative control outcomes were utilized for empirical calibration of results. (eFigure 5) The proportion of 95% confidence intervals for negative control estimates that included the null was 96.2% in HIRA, 94.0% in HealthVerity, and 95.5% in CCAE after calibration and minimal residual systematic error was achieved.

Findings were consistent across prespecified sensitivity analyses (Figure 4). Under the as-treated definition, the pooled HR was 1.41 (95% CrI, 0.95–2.09). For alternative fixed follow-up windows, pooled HRs were 1.37 (95% CrI, 0.95–2.03) at 3 months and 1.27 (95% CrI, 0.82–2.02) at 1 month. Across sensitivity analyses, PS-stratified point estimates were directionally toward favoring prasugrel but were accompanied by wide credible intervals, thereby not resulting in statistically credible pooled differences. In contrast, PS-matched analyses yielded more attenuated estimates, including a pooled HR of 1.12 (95% CrI, 0.82– 1.57) for the 1-year time-at-risk (Figure 4).

### Secondary outcomes

Pooled estimates for 1-year secondary outcomes are shown in Figure 3. In Bayesian random-effects meta-analyses, none of the secondary endpoints demonstrated a statistically credible difference between ticagrelor and prasugrel.

**Figure 3.**
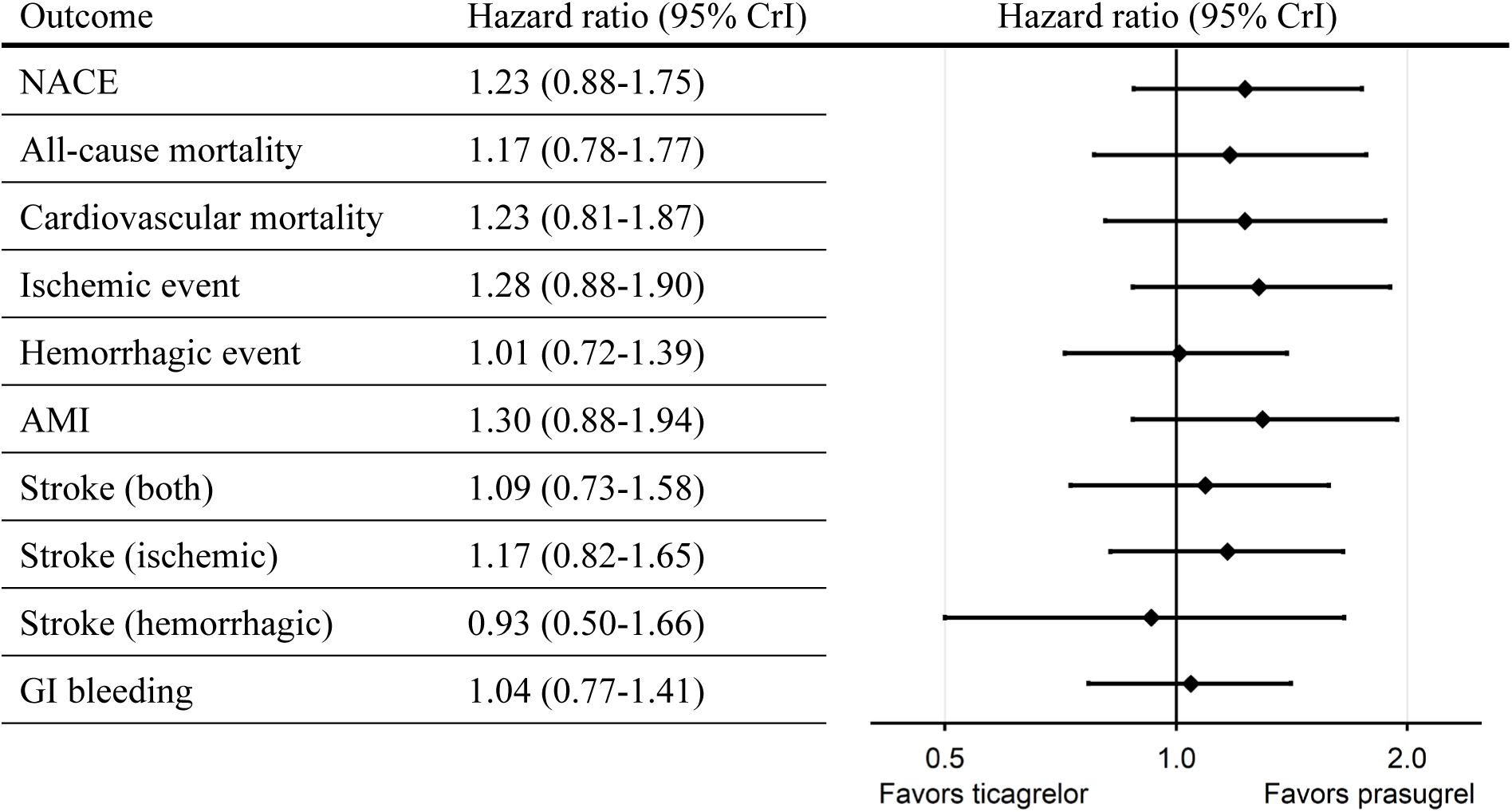
Meta-analysis of 1-year secondary outcomes. Forest plot of pooled hazard ratios (HRs) and 95% credible intervals (CrIs) for 1-year secondary clinical outcomes comparing ticagrelor with prasugrel. Estimates were derived using Bayesian random-effects meta-analysis. Points represent posterior mean HRs, and horizontal lines indicate 95% CrIs. The vertical line denotes no difference (HR = 1.0). HRs greater than 1.0 favor prasugrel over ticagrelor. Abbreviations: CrI, credible interval; NACE, net adverse clinical events; AMI, acute myocardial infarction; GI, gastrointestinal.

**Figure 4.**
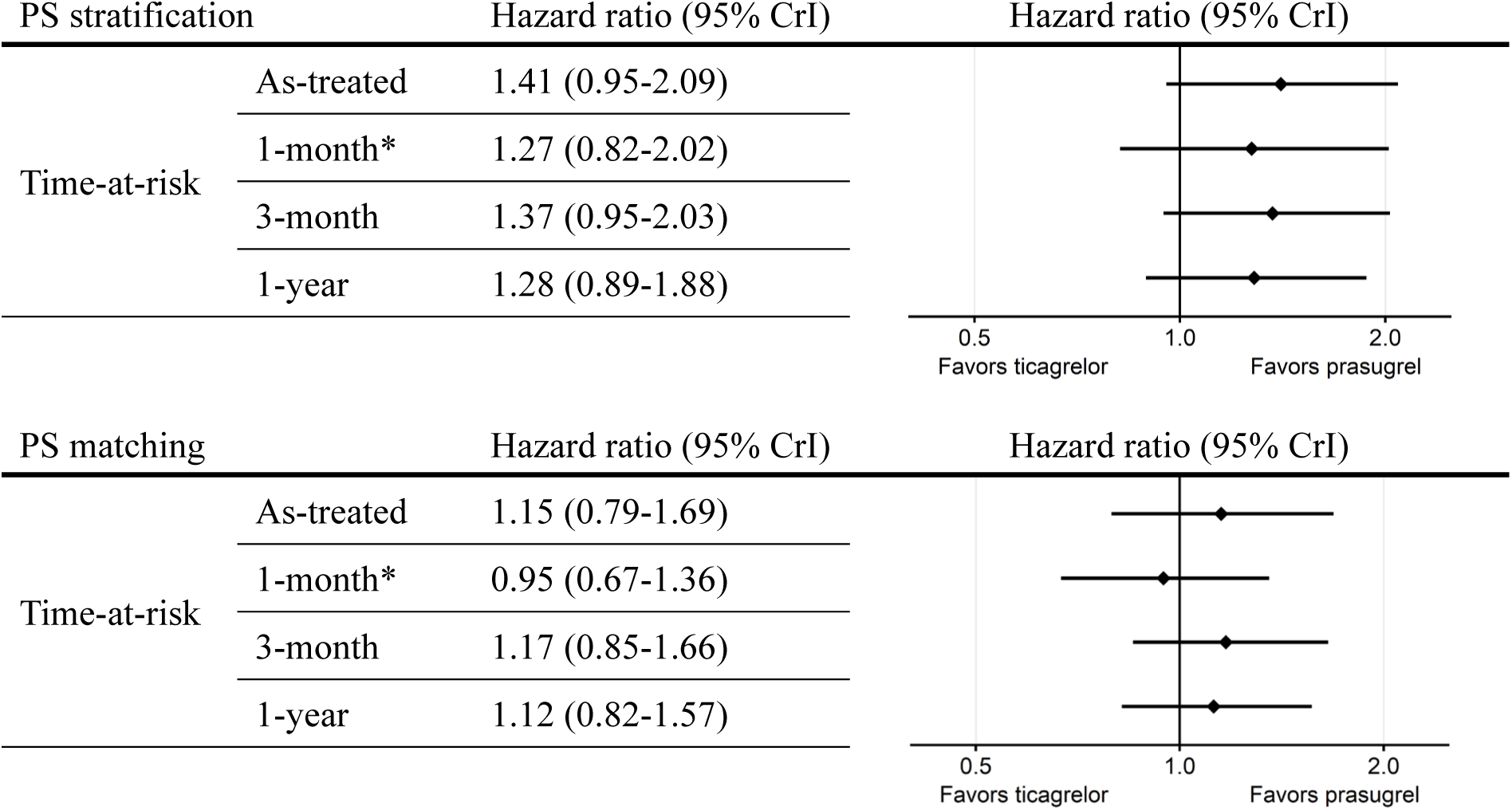
Sensitivity analyses of major adverse cardiovascular events. Forest plots of pooled hazard ratios (HRs) and 95% credible intervals (CrIs) for sensitivity analyses of major adverse cardiovascular events (MACE) comparing ticagrelor with prasugrel across alternative time-at-risk definitions (1-year, 3-month, 1-month, and as-treated) and propensity score (PS) adjustment strategies (PS stratification and PS matching). Estimates were derived using Bayesian random-effects meta-analysis. Points represent posterior mean HRs, and horizontal lines indicate 95% CrIs. The vertical line denotes no difference (HR = 1.0). HRs greater than 1.0 favor prasugrel over ticagrelor. *The 1-month analysis includes data from HIRA and HealthVerity only; CCAE was excluded because prespecified study diagnostics were not met for this time-at-risk definition. Abbreviations: PS, propensity score; CrI, credible interval.

For net adverse clinical events (NACE), the pooled HR was 1.23 (95% CrI, 0.88–1.75). For all-cause mortality, the pooled HR was 1.17 (95% CrI, 0.78–1.77), and for cardiovascular mortality, 1.23 (95% CrI, 0.81–1.87). For ischemic outcomes, the pooled HR was 1.28 (95% CrI, 0.88–1.90) for the composite ischemic endpoint and 1.30 (95% CrI, 0.88–1.94) for acute myocardial infarction. Stroke outcomes were similarly neutral, with pooled HRs of 1.09 (95% CrI, 0.73–1.58) for any stroke, 1.17 (95% CrI, 0.82–1.65) for ischemic stroke, and 0.93 (95% CrI, 0.50–1.66) for hemorrhagic stroke. For bleeding-related outcomes, the pooled HR was 1.01 (95% CrI, 0.72–1.39) for the composite hemorrhagic endpoint and 1.04 (95% CrI, 0.77– 1.41) for gastrointestinal bleeding.

Sensitivity analyses of secondary outcomes across alternative time-at-risk definitions and PS adjustment strategies are shown in eFigure 7. Results were consistent with the primary 1-year analyses and did not demonstrate statistically credible differences.

### Post-hoc analyses

Given the substantial heterogeneity observed in the primary multinational analysis, a post hoc meta-analysis restricted to U.S. databases (HealthVerity and CCAE) was conducted (Figure 5).

**Figure 5.**
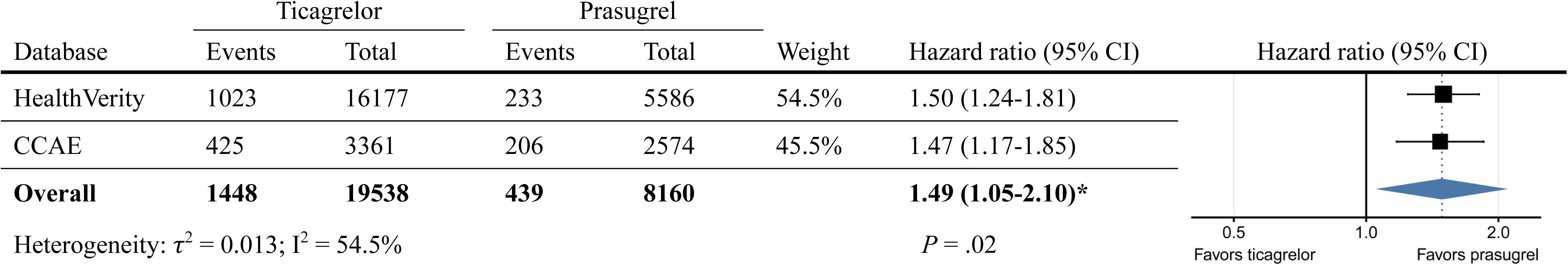
Post hoc U.S.-only meta-analysis of 1-year major adverse cardiovascular events. Forest plot of calibrated hazard ratios (HRs) for 1-year major adverse cardiovascular events restricted to U.S. databases (HealthVerity and CCAE). Database-specific HRs and 95% confidence intervals (CIs) were estimated using propensity score-stratified Cox proportional hazards models, and were empirically calibrated. The pooled HR was obtained using Bayesian random-effects meta-analysis and is presented with its 95% credible interval (CrI). Weights are normalized random-effects inverse-variance weights based on the posterior median estimate of between-database heterogeneity. Square sizes are proportional to the weights, and horizontal lines indicate 95% CIs for database-specific estimates. The diamond represents the pooled posterior mean HR with its 95% CrI. The vertical line denotes no difference (HR = 1.0). HRs greater than 1.0 favor prasugrel over ticagrelor. Between-database heterogeneity is summarized by τ², the estimated between-database variance in the true log HRs, and I², the proportion of total variability attributable to between-database heterogeneity. *P* represents the P value for the overall pooled effect. *The pooled estimate is derived from Bayesian random-effects meta-analysis; the interval shown represents a 95% credible interval (CrI). Abbreviations: CI, confidence interval; CCAE, Merative MarketScan Commercial Claims and Encounters database.

The pooled Bayesian random-effects estimate yielded an HR of 1.49 (95% CrI, 1.05– 2.10), indicating a statistically credible association favoring prasugrel in the U.S. population (Figure 5). Between-database heterogeneity was moderate (τ² = 0.013; I² = 54.5%). For secondary outcomes in the U.S.-only meta-analysis, the composite ischemic endpoint and acute myocardial infarction (AMI) demonstrated statistically credible associations favoring prasugrel. (eFigure 8) In contrast, other secondary endpoints including NACE, mortality outcomes, stroke subtypes, and bleeding outcomes had credible intervals crossing 1.

Sensitivity analyses restricted to U.S. databases are shown in eFigure 9. Under PS stratification, results were consistent across alternative time-at-risk definitions, with statistically credible associations favoring prasugrel. In contrast, PS-matched analyses yielded attenuated estimates, and credible intervals crossed 1 for all time-at-risk definitions. These findings indicate that the observed U.S.-specific signal was robust to alternate follow-up definitions under PS stratification but sensitive to the adjustment approach.

In the separate exploratory analysis using expanded cohort definitions, both CCAE and MDCR met all prespecified diagnostic criteria (eTable 6). The calibrated HRs for 1-year MACE were 1.25 (95% CI, 1.02–1.54) in CCAE and 1.43 (95% CI, 1.13–1.80) in MDCR. When these estimates were combined with the HIRA and HealthVerity estimates derived using the original cohort definition, the pooled HR was 1.26 (95% CrI, 0.97–1.66). In the corresponding U.S.-only meta-analysis, the pooled HR was 1.39 (95% CrI, 1.10–1.76), favoring prasugrel (eFigure 10). These findings were consistent with the main analyses, showing no statistically credible overall difference but a prasugrel-favoring association in the U.S.-only analysis. Because different cohort definitions were applied across databases, these findings were considered exploratory.

## DISCUSSION

In this multinational observational cohort study, we observed no statistically credible overall difference in 1-year MACE between ticagrelor and prasugrel. However, effect estimates varied substantially across databases, with neutral associations in the nationwide Korean database and higher hazards with ticagrelor in the U.S. databases. Results were generally consistent across sensitivity analyses, and analyses of 100 negative control outcomes indicated limited residual systematic error. A post hoc exploratory analysis using expanded cohort definitions in CCAE and MDCR showed the same overall pattern: no statistically credible difference in the pooled estimate but a prasugrel-favoring association in the U.S.-only analysis. The overall pooled neutrality is broadly consistent with results from PRAGUE-18, although that trial was limited by early termination.5,^6^ Conversely, the statistically credible associations observed in the U.S.-only post hoc analysis favoring prasugrel are directionally aligned with ISAR-REACT 5, which demonstrated lower ischemic event rates with prasugrel without an increase in major bleeding.^7^ The recent TUXEDO-2 trial, which failed to prove noninferiority of ticagrelor in a high-risk diabetic PCI population also aligns with this association observed in our post-hoc analysis.^11^ However, differences in study populations and designs, together with the post hoc nature of our U.S.-only analysis, limit direct comparisons among these findings.

This study extends prior observational evidence by applying a single prespecified analytic framework, common cohort and outcome definitions, standardized diagnostics, and empirical calibration across independent healthcare systems. Previous observational studies have produced mixed estimates. Some U.S. claims-based analyses favored prasugrel for ischemic outcomes without clear excess major bleeding,19,^21^ whereas another reported ticagrelor-favoring ischemic and bleeding outcomes.^43^ European multicenter registry data have reported prasugrel-favoring signals in selected ACS presentations,^44^ while nationwide Scandinavian registry data and large Korean population-based studies have generally shown neutral comparative effectiveness.^18,45,46^ Because these studies differed in their populations, data sources, cohort definitions, and analytic methods, contextual variation could not be readily distinguished from methodological variation. In our study, substantial heterogeneity persisted despite harmonized design and execution, indicating that discrepancies among real-world estimates cannot be attributed solely to analytic specifications and may also reflect differences in patient populations, treatment selection, and healthcare context.

The absence of a statistically credible difference in our overall meta-analysis mainly stems from divergence in effect estimates across regions, which yielded wider posterior uncertainty in the pooled effect. The nationwide Korean HIRA database, which contributed the largest proportion of patients, demonstrated largely neutral results across both ischemic and hemorrhagic outcomes. In contrast, the U.S. databases showed higher hazards of the primary endpoint with ticagrelor, primarily driven by ischemic outcomes. One plausible explanation for this regional divergence is differences in underlying population composition: HIRA largely represents an East Asian population, whereas the race data available in HealthVerity suggests a predominantly White population. Racial differences in pharmacokinetic exposure and pharmacodynamic platelet inhibition with potent P2Y₁₂ inhibitors have been reported, providing a biologically plausible mechanism by which comparative effects could vary across populations.47,^48^ In addition, health-system and socioeconomic factors such as insurance design and out-of-pocket costs can influence initiation and adherence with P2Y₁₂ inhibitors after PCI and shape observed comparative outcomes in routine care.49,^50^ However, because population, database, and healthcare-system characteristics were inseparable in this study, the observed heterogeneity cannot be attributed to any specific factor alone. Disentangling these influences will require studies designed to evaluate specific determinants of comparative effectiveness.

This study has several strengths. First, analyses were conducted according to a prespecified, publicly registered analytic protocol developed prior to study execution, enhancing transparency and minimizing analytic variability. Second, implementation within the OHDSI framework using the OMOP-CDM enabled standardized cohort definitions, outcome algorithms, and analytic procedures across independent international databases, enhancing reproducibility. Third, we applied large-scale propensity score adjustment incorporating extensive baseline covariates and required stringent prespecified study diagnostics before inclusion in the meta-analysis, thereby reducing confounding. Fourth, empirical calibration using negative control outcomes, and sensitivity analyses across alternative propensity score adjustment strategies and time-at-risk definitions further supported the robustness of findings. Finally, the study leveraged large-scale real-world data from multiple healthcare systems of different countries, including the nationwide HIRA database. HIRA captures healthcare utilization across all levels of care in South Korea, contributing to population representativeness.

Several limitations warrant consideration. First, as with all observational studies, residual confounding, including confounding by indication or treatment selection, cannot be completely excluded despite extensive propensity score adjustment, prespecified diagnostics, and empirical calibration. Second, although individual outcomes were separately validated through chart review, code-based algorithms remain susceptible to coding errors and misclassification. Moreover, detailed outcome definition incorporating established bleeding classifications could not be performed and therefore relied on coded diagnosis of bleeding. Third, because claims databases lack granular clinical detail, important factors such as lesion complexity and procedural characteristics were unavailable. In addition, information not fully captured in prescription data—including medication adherence and over-the-counter aspirin use—could not be assessed. Furthermore, some variables were incompletely captured due to database-specific characteristics. Race information was unavailable in HIRA and CCAE, precluding formal assessment of effect modification by race. U.S. claims databases such as CCAE may also incompletely capture out-of-hospital mortality, potentially affecting outcome ascertainment.^51^ Fourth, within the OMOP-CDM-based study framework, detailed stratification or longitudinal assessment of P2Y₁₂ inhibitor dosing, as well as concomitant aspirin dosage, could not be performed, which may have influenced the observed outcomes. Fifth, only three databases met prespecified diagnostic criteria and contributed to the meta-analysis. Although this approach strengthened the validity of our results, broader database participation might have provided additional insight into the observed heterogeneity.

In conclusion, our multinational analysis utilizing the OHDSI network did not identify a clear overall advantage of ticagrelor versus prasugrel for 1-year MACE, which was consistent across secondary endpoints and sensitivity analyses. At the same time, high cross-database heterogeneity was observed, with neutral estimates in nationwide Korean claims database and prasugrel-favoring ischemic signals in U.S. claims databases. These findings suggest that comparative effectiveness may not be uniform across populations and health-care settings, and caution against extrapolating comparative-effectiveness estimates from one care context to another.

## Data Availability

The patient-level data used in this study cannot be made publicly available. Additional aggregated study results may be available from the corresponding author upon reasonable request, subject to approval by the participating data partners and applicable data-use restrictions. Analytic code used for this study is publicly available through the study repositories cited in the manuscript.

## ACKNOWLEDGMENTS

This study used HIRA OMOP-CDM data made by Health Insurance Review & Assessment Service(HIRA). The views expressed are those of the author(s) and not necessarily those of the HIRA and the Ministry of Health & Welfare, Republic of Korea.

## TABLES

